# Excess deaths during the Covid-19 pandemic: An international comparison

**DOI:** 10.1101/2020.04.21.20073114

**Authors:** Kieran F. Docherty, Jawad H. Butt, Rudolf A. de Boer, Pooja Dewan, Lars Køber, Aldo P. Maggioni, John J.V. McMurray, Scott D. Solomon, Pardeep S. Jhund

## Abstract

**Background:** With the global pandemic of coronavirus disease 2019 (Covid-19) there has been disruption to normal clinical activity in response to the increased demand on health services. There are reports of a reduction in non-Covid-19 emergency presentations. Consequentially, there are concerns that deaths from non-Covid-19 causes could increase. We examined recent reported population-based mortality rates, compared with expected rates, and compared any excess in deaths with the number of deaths attributed to Covid-19.

**Methods:** National agency and death registration reports were searched for numbers of deaths attributed to Covid-19 and overall mortality that had been publicly reported by 06 May 2020. Data on the number of deaths attributed to Covid-19, the total number of deaths registered in the population and the historical average over at least 3 years were collected. Data were available for 4 European countries (England & Wales, Scotland, Netherlands and Italy) and New York State, United States of America.

**Results:** There was an increase in observed, compared with expected, mortality in Scotland (+68%), England and Wales (+74%), the Netherlands (+58%), Italy (+39%) and New York state (+49%). Of these deaths, only 73% in Scotland, 71 % in England and Wales, 53% in the Netherlands, 54% in Italy and 79% in New York state were attributed to Covid-19 leaving a number of excess deaths not attributed to Covid-19. In the 5-week period of study, Scotland, 10% of the excess of deaths were attributed to dementia/Alzheimer’s disease and 7% to cardiovascular causes.

**Conclusion:** A substantial proportion of excess deaths observed during the current COVID-19 pandemic are not attributed to COVID-19 and may represent unrecognised deaths due to Covid-19, an excess of deaths due to other causes, or both. The impact of Covid-19 on mortality and morbidity from other causes needs to be quantified and addressed in public health planning.

## INTRODUCTION

Since the first report of a novel pneumonic illness in Wuhan, China, in December 2019, the severe acute respiratory syndrome coronavirus 2 (SARS-Cov-2) has spread globally causing a pandemic that, at the time of writing, has infected approximately 3.7 million people and resulted in over 260,000 deaths attributed to the illness it causes, coronavirus disease 2019 (Covid-19).^1,2^Most official reports of Covid-19-related mortality are based on hospital data with variable inclusion of deaths in the community, possibly resulting in under-recognition of the full impact of SARS-Cov-2 on population death rates. The huge influx of Covid-19 patients into hospitals in affected countries has led to disruption of usual clinical activity, with cancellation of elective in-patient and outpatient activity.^3-5^ In many cases, normal ambulatory care has been replaced by remote management using a variety of telemedicine approaches.^6^ Even non-Covid-19 related emergency department activity has decreased markedly in many countries, as reported in a previous Severe Acute Respiratory Syndrome outbreak.^7,8^ As a result, there is concern that deaths from non-Covid-19 causes could increase, due to reduced routine care. One way to address these two questions is to examine recent observed population-based mortality rates, compared with expected rates, and compare any excess in deaths with the number of deaths attributed to Covid-19. We have analysed these data for 5 countries which have provided such data on a routine basis for at least 5 years.

## Methods

We examined recent changes in population mortality following the outbreak of Covid-19 in Northern European Countries with similar population characteristics, universal health care systems and rapid and complete reporting of data on deaths. The countries identified also had outbreaks of Covid-19 starting at approximately the same time. We included the constituent countries of the United Kingdom which collect, collate and report national data separately (for the purposes of this report, England and Wales were considered one country as their data are reported together) and the Netherlands in this analysis. The first case of Covid-19 was reported England 31 January and the first death 5 March 2020. These dates were 27 February/6 March for the Netherlands, 28 February/16 March for Wales and 1 March/13 March for Scotland. In addition, we examined mortality in Italy, one of the worst and earliest affected countries in Europe. The first case of Covid-19 reported in Italy was 31^st^ January 2020 and the first death recorded on 22^nd^ February 2020.

The populations of the countries studied were: Scotland 5,438,100, England and Wales 59,115,809, Italy 60,359,546 and the Netherlands 17,213,114, based on published 2018 mid-year estimates. We examined data from New York State (and New York city) for the period 26 January 2020 to April 18 2020 (state population 19,542,209, city population 8,398,748). The first case of Covid-19 was reported in New York State 1 March and the first death 14 March 2020.As this analysis utilised publicly available national statistics, no ethical approval was required.

For the European countries included and New York, we extracted reports of the numbers of deaths related to Covid-19 published by national agencies and death registration authorities that had been publicly reported by 6^th^ May 2020.^9-13^ Data on the number of deaths due to Covid-19, and the total number of deaths registered in the population over the same period were collected. Historical numbers of deaths or average number of deaths from 2015-2019 for the same week were extracted from available records (data were available only for 2015-2018 for New York State and 2015-2017 for New York City). The number of excess deaths was defined as the difference between the observed number of deaths and the number of deaths expected according to the observed number from 2015-2019. The number of deaths related to Covid-19 was then deducted from the excess deaths to calculate the number of non-Covid-19 defined excess deaths. Numbers of deaths were also converted into a rates per million population using 2018 mid-year population estimates and 95% confidence intervals computed. The underlying or main cause of death was reported for Scotland for the period 30th March – 3rd May 2020. The underlying causes of death for the excess deaths is expressed as a number and rate per million population per 7 days. England and Wales was the only country to report age specific deaths and in a subgroup analysis we explored the relationship between age and excess deaths in England and Wales. Deaths were categorised according to the following age groups, <45, 45-64, 65-74, 75-84 and ≥85. These categories were used as historical data on deaths was available in these age categories. All analyses were carried out using Stata software, version 16 (StataCorp, College Station, TX, USA).

## RESULTS

### United Kingdom

#### Scotland

There were 3715 excess deaths in Scotland in the period 30 March to 3 May 2020 compared to the same period in 2015-2019 (an increase of 68%). Of these excess deaths only 2723 (73%) were recorded as related to Covid-19 (Figure 1). As a result, the death rate for the 3-week period examined rose from an expected 200 per million (95% confidence interval [CI] 195 to 205), based on the 5-year average, to 336 per million of population (95% CI 330 to 343). The 7 day rate of excess deaths attributed to Covid-19 was 100 per million (95% CI 96 to 104), and the rate of excess deaths not attributed to Covid-19 was 36 per million per 7 days (95% CI 34 to 39).

**Figure 1.**
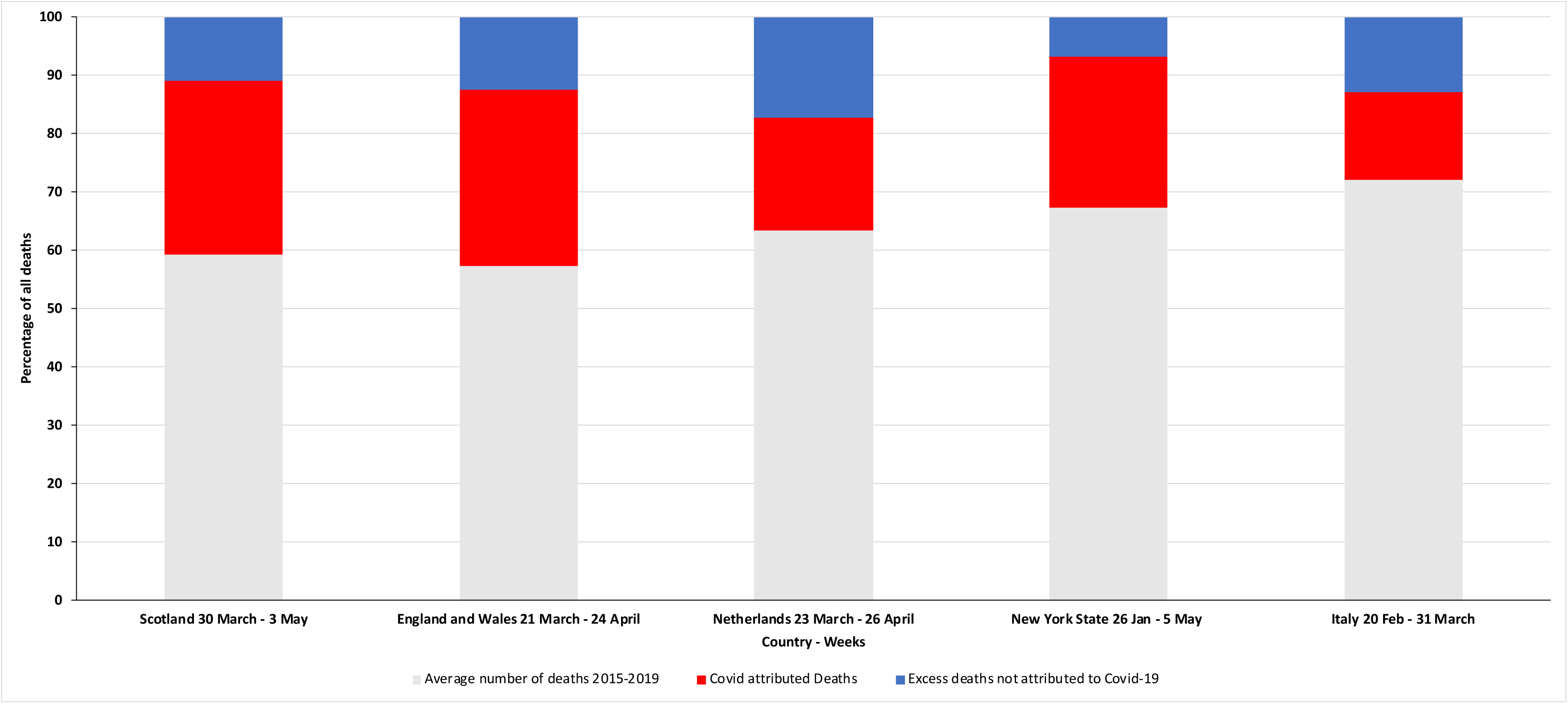
Deaths in Scotland, England and Wales, the Netherlands and New York State during the Covid-19 Pandemic. Total deaths observed in each country are normalised to 100%. Expected deaths, as a percentage of total deaths observed, are shown in grey. Excess deaths attributed to Covid-19 are shown in red and excess deaths not attributed to Covid-19 are shown in blue.

#### England and Wales

In England and Wales there were 38482 excess deaths in the period 21 March to 24 April 2020, a 74% increase from the prior 5-year average. Of these excess deaths, 27222 (71%) were Covid-19-related. As a result, the death rate rose from an expected of 176 per million per 7 days (95% CI 174 to 177) to 306 (95% CI 304 to 308). The rate of Covid-19-related deaths was 92 per million per 7 days (95% CI 91 to 93), and the rate of excess deaths not attributed to Covid-19 was 38 (95% CI 37 to 39).

### Italy

In Italy the number of deaths increased from an expected 65592 to 90946 (39% increase) over the period 20 February to 31 March 2020. Of the excess deaths recorded, 13710 were ascribed to Covid-19 deaths. The overall death rate during this period rose from an expected 186 per million per 7 days (95%CI 184-187) to 257 (95%CI 256-259). The rate of excess Covid-19 deaths was 39 per million per 7 days (95%CI 38-39) and the rate of excess deaths not attributed to Covid-19 was 33 (95%CI 32-33).

### Netherlands

In the Netherlands the number of deaths increased from an expected 14423 (based on the prior 5-year average) to 22761 (58% increase) over the period 23 March to 26 April 2020. Of the 8338 excess deaths, 4399 (52%) were related to Covid-19. As a result, the death rate rose from an expected 168 per million per 7 days (95% CI 165 to 170) to 264 (95% CI 261 to 268). The rate of Covid-19-related deaths was 51 per million per 7 days (95% CI 50 to 53), and the rate of excess deaths not attributed to Covid-19 was 46 (95% CI 44 to 47).

### New York State

In the New York state the number of deaths increased from an expected 44894 (based on the prior average from 2015-2018) to 66771 (49% increase) over the period 26 January to 5 May 2020. Of the 21877 excess deaths, 17356 (79%) were related to Covid-19. As a result, the overall death rate for the period rose from an expected 159 per million per 7 days (95% CI 58 to 161) to 237 (95% CI 235 to 239). The rate of Covid-19-related deaths was 62 per million per 7 days (95% CI 61 to 62), and the rate of excess deaths not attributed to Covid-19 was 16 (95% CI 16 to 16). The equivalent figures for New York city were a 112% increase from 15618 expected deaths (based on 2015-2017 average) to 33100 deaths over the period 26 January to 5 May 2020. Of the 17482 excess deaths, 12178 (70%) were related to Covid-19. As a result, the overall death rate rose from an expected 129 per million per 7 days (95% CI 127 to 131) to 273 (95% CI 270 to 276). The rate of Covid-19-related deaths was 100 per million per 7 days (95% CI 99 to 102), and the rate of excess deaths not attributed to Covid-19 was 44 (95% CI 43 to 45).

### Age specific excess deaths in England and Wales

In those <45 years there were 1978 reported deaths compared to an average of 1789. Of these 1978 deaths 331 (16.7%) were attributed to Covid-19, with no clear excess of non-Covid-19 deaths. In all other age groups, there was a clear excess of both deaths attributed to Covid-19 and deaths not attributed to Covid-19 (Figure 2). In those aged 45-64 there were 9849 deaths compared to an average of 6091. Of these excess deaths Covid-19 accounted for 3006 deaths (80%). A similar pattern was seen for those aged 65-74 years with 14103 deaths reported compared to a historical average of 8527. Of the excess deaths, 4460 (80%) were attributed to Covid-19. In those aged 75-84 years the respective figures were 27035 total deaths compared to 14733 historical deaths, with 9071 Covid-19 deaths accounting for 74% of the excess deaths. In the eldest group (≥ 85 years) there were 37428 deaths recorded compared to an average of 20749. There was still an excess of deaths but only 10354 deaths (62%) were attributed to Covid-19.

**Figure 2.**
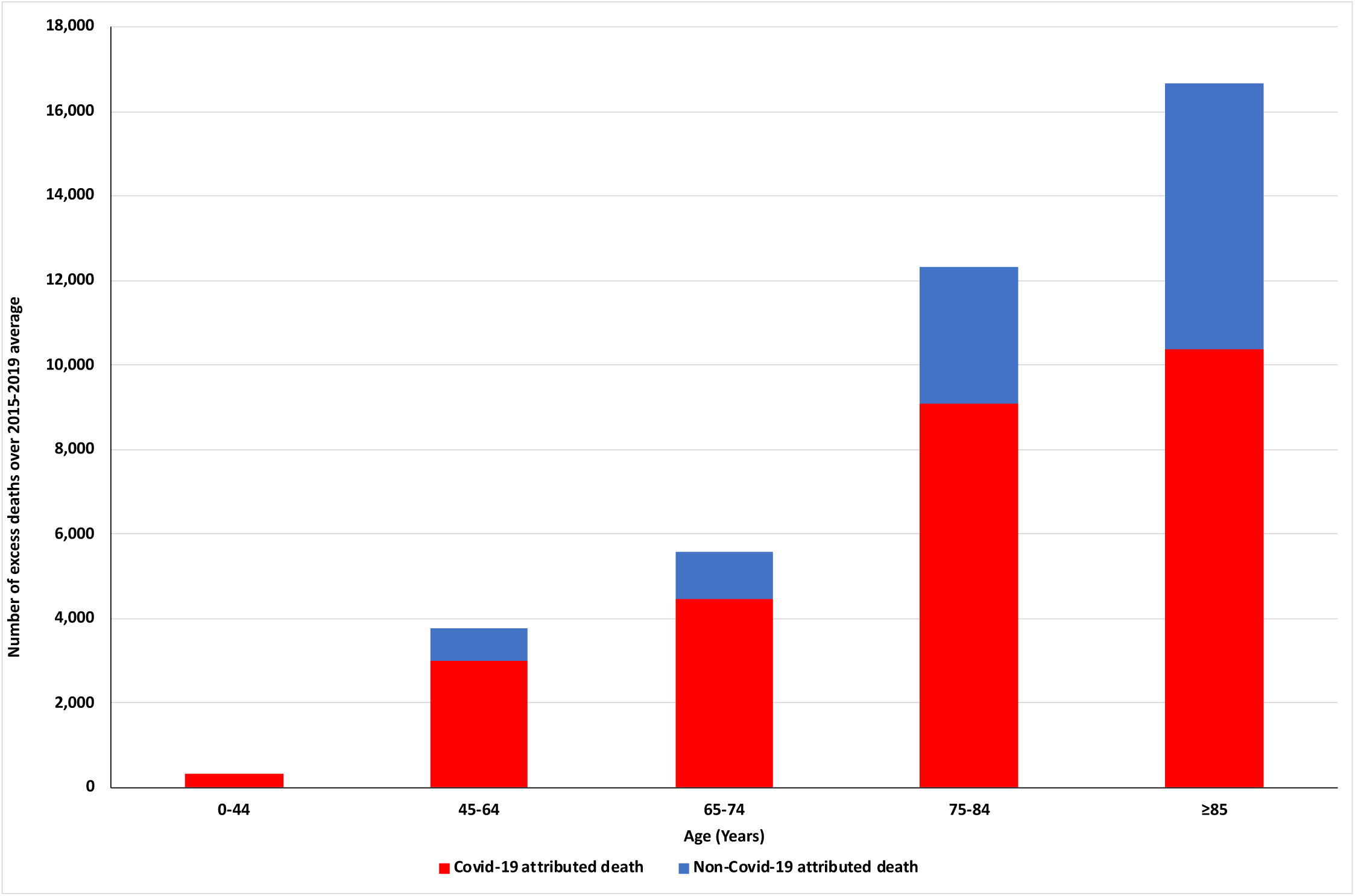
Excess Deaths above the 2015-2019 average in England and Wales by age group during the Covid-19 Pandemic. Deaths are those registered between 21 March and 24 April 2020 compared to the 2015-2019 average during the same time-period. The number of excess deaths attributed to Covid-19 are shown in red and excess deaths not attributed to Covid-19 are shown in blue.

### Cause of excess deaths in Scotland

For the period 30 March to 3 May 2020 data on the underlying (main cause) of death were available for Scotland. Figure 3 shows the percentage of excess deaths by cause. The majority of excess deaths were due to Covid-19 (the proportions being 39%, 67%, 75% 85% and 83% in the weeks beginning 30 March, 13 April, 20 Apr and 27 April respectively). A significant proportion of the non-Covid-19 excess deaths were due to dementia/Alzheimer’s disease (International Classification of Diseases 10th revision [ICD] codes F01, F03, G30), accounting for 10% of deaths over the 5-week period. Deaths from circulatory causes (ICD codes I00-99) accounted for an additional 7% of the excess over the 5 weeks. Conversely, cancer deaths (ICD codes C00-C97) fell over the 5 weeks, so that in the most recent two weeks there were fewer deaths from cancer than expected. Non-Covid-19-related respiratory deaths (ICD codes J00-J99) were also fewer than expected in the latter part of the 5-week period.

**Figure 3.**
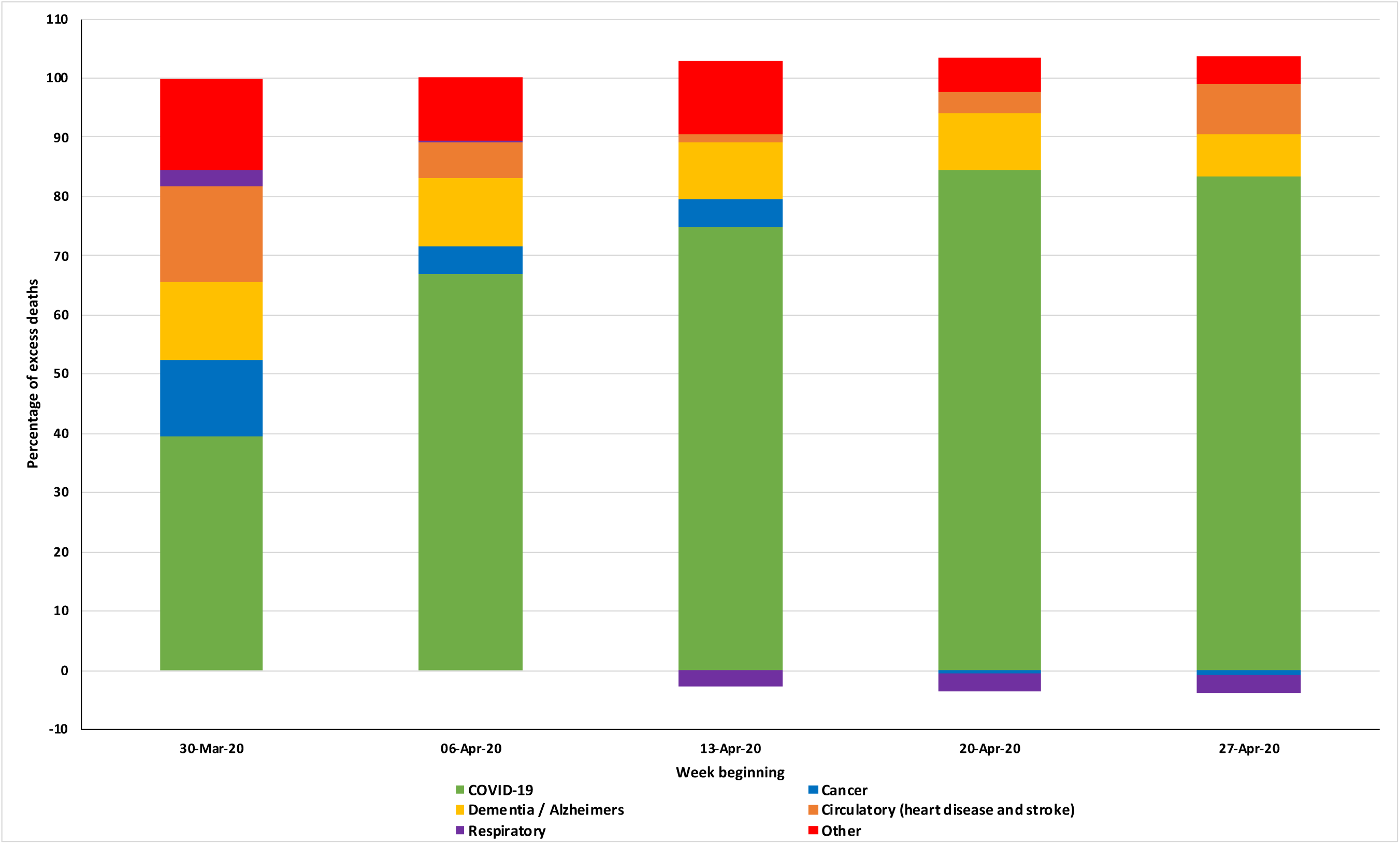
Excess deaths by underlying cause on the death certificate registered between 30 March to 3 May 2020 in Scotland. Proportions may add to more than 100% due to less than expected deaths for a particular category compared to the previous year.

## DISCUSSION

In the countries analysed, there has been a substantial increase (ranging from 39 to 75%) in observed, compared with expected, mortality at a population level. The increase in mortality is not wholly explained by deaths attributed to Covid-19. Indeed, only between 53 and 80% of the excess in deaths can be explained by official Covid-19 reports. The data from New York state support these findings, with an even more striking picture in New York city. The two most likely explanations for the discrepancy between the overall excess of deaths and the extra deaths not explained by Covid-19 are either there are additional deaths caused (or contributed to) by Covid-19, but not recognised as such, or that there is an increase in deaths from non-Covid-19 causes, potentially resulting from diminished routine diagnosis and treatment of other conditions. We believe that both are likely. In many reports of mortality attributed to Covid-19, the focus has been on deaths occurring in hospital or in those with a confirmed diagnosis by testing. Similarly, in some countries due to limited testing for SARS-Cov-2 infection, deaths among elderly people in care homes and other places potentially caused (or contributed to) by Covid-19 are not attributed to that condition.^14^ The excess deaths was attributed to dementia/ Alzheimer’s disease in Scotland support this possibility. In keeping with this, the proportion of excess deaths attributed to Covid-19 was smallest in the oldest age group in England and Wales. More frequent comorbidity in older individuals may make the cause of death less certain or make it easier to ascribe an alternative cause of death.

However, the extra cardiovascular deaths reported in Scotland suggest the recent excess mortality rate may not just be due to unrecognised Covid-19 deaths. There are anecdotal reports of people with Covid-19-like symptoms frightened to attend hospital or choosing to remain at home because of fear of dying in isolation from their family in hospital^15^. The latter behaviours may also be relevant to deaths from non-Covid-19 causes. Not only has elective clinical activity almost ceased in many countries but emergency presentations have also declined markedly ^8^ and for some conditions, for example acute coronary syndromes, this could have fatal consequences.^5,16,17^Rates of primary percutaneous coronary intervention (PPCI) have declined and even where PPCI services have continued, disruption of normal working, including staff shortages due to illness or redeployment, and even donning personal protection equipment may have lengthened time to reperfusion. Even return to use of thrombolysis has been mooted.^18^ In Italy where we have documented these excess deaths, a reduction in rates of treatment for acute coronary syndromes ^17^ has been accompanied by an increase in the expected numbers of out of hospital cardiac arrests^19^. The same considerations apply to acute stroke.^20^ There has also been concern about re-supply of essential medicines to patients and the impact of public discussion of the safety ACE inhibitors and ARBs, which are life-saving treatments in patients with heart failure and reduced ejection fraction and in high-risk survivors of myocardial infarction, is difficult to quantify.^21^

Similarly, the excess of “other” deaths is of interest. Mental health problems and domestic violence may also add to the non-Covid-19 excess in mortality, although reduction in industrial deaths, fatal road traffic accidents and deaths related to air pollution may offset increases from these other causes.^22,23^ Clearly, therefore, the causes of death in the “other” category will require further examination, especially by age, when that information becomes available.

Clearly it will take time to fully explain the trends we have described and, especially, to quantify the exact causes of the excess of non-Covid-19 deaths, although even with more data, it may be difficult to accurately determine whether Covid-19 caused or contributed to death. However, if it is correct that the Covid-19 pandemic has had a detrimental effect on medical care more generally, other “downstream” consequences are likely^24^. Non-Covid-19 hospital admissions may rebound as chronic conditions destabilise. Failure to diagnosis and treat new events may have other delayed consequences, for example the development of heart failure in sub-optimally treated or untreated acute myocardial infarction and stroke in patients with untreated transient ischaemic attack. The consequences for conditions with less acute presentations, such as cancer, may take longer to become fully apparent.

What are the implications of these findings and why is it important to highlight them now rather than wait for the more granular information that will become available in time? We believe that the public health message to “stay at home” during period of social distancing needs to be more nuanced, with an equally clear message to patients with symptoms suggestive of an acute coronary syndrome, stroke and worsening of chronic heart failure to still seek urgent medical attention. This may need reconfiguration of admission procedures to avoid mixing of suspected Covid-19 patients and other emergencies. Similarly, our findings reinforce the necessity of efforts being made to adapt ambulatory secondary care, and primary care, services to enable the continued delivery of essential treatment to high risk patients with chronic diseases and the triage of newly presenting individuals with potentially life-threatening conditions.

Similarly, there is a need to expedite and standardise reporting of mortality during the pandemic. If there is an excess of non-Covid-19 deaths and non-fatal events due to inaccessible or suboptimal healthcare during the pandemic, the effects on long-term total mortality (and morbidity) may be far-reaching and strategies to detect and prevent this must be developed and shared. Sustained monitoring will also be required to demonstrate the abatement of any such impact.

Our analysis has several limitations. It would have been of interest to include other countries but we could only use those providing the required data. We relied on publicly available data that are collected and collated in a fast-moving pandemic and may be subject to revision. The coding and therefore definition of a Covid-19 related death may be influenced by local regulations and guidelines on certifying the cause of death in each country, as discussed in relation to Spain above. Finally, the availability of testing for Covid-19 may have influenced the ascertainment of Covid-19 related deaths in each country and therefore the excess of non-Covid-19 deaths in each of the countries may represent an under-identification of cases of SARS-Cov-2 infection. Conversely, the number of Covid-19 deaths may be inflated by deaths in people who had Covid-19 at the time of death but did not die from their viral illness i.e. died from another non-Covid-19 reason despite infection with SARS-Cov-2. The lack of age specific data in countries other than England and Wales prevented the calculation of age adjusted rates and the ability to compare using standardised mortality statistics.

In summary, a substantial proportion of excess deaths observed during the current COVID-19 pandemic are not attributed to COVID-19. This may indicate an increase in non-COVID-19 deaths due to changes in routine health care delivery during this pandemic. People should be reminded that it is still appropriate to seek medical attention for other serious life-threatening illnesses during this period and service delivery adapted to provide this. Plans for future waves of the pandemic must account for these excess deaths.

## Data Availability

All data are publicly available

https://www.nrscotland.gov.uk/covid19stats

https://www.ons.gov.uk/peoplepopulationandcommunity/birthsdeathsandmarriages/deaths/datasets/weeklyprovisionalfiguresondeathsregisteredinenglandandwales

https://www.rivm.nl/coronavirus-covid-19/grafieken

https://www.cdc.gov/nchs/nvss/vsrr/COVID19/index.htm

---

**Table 1:**
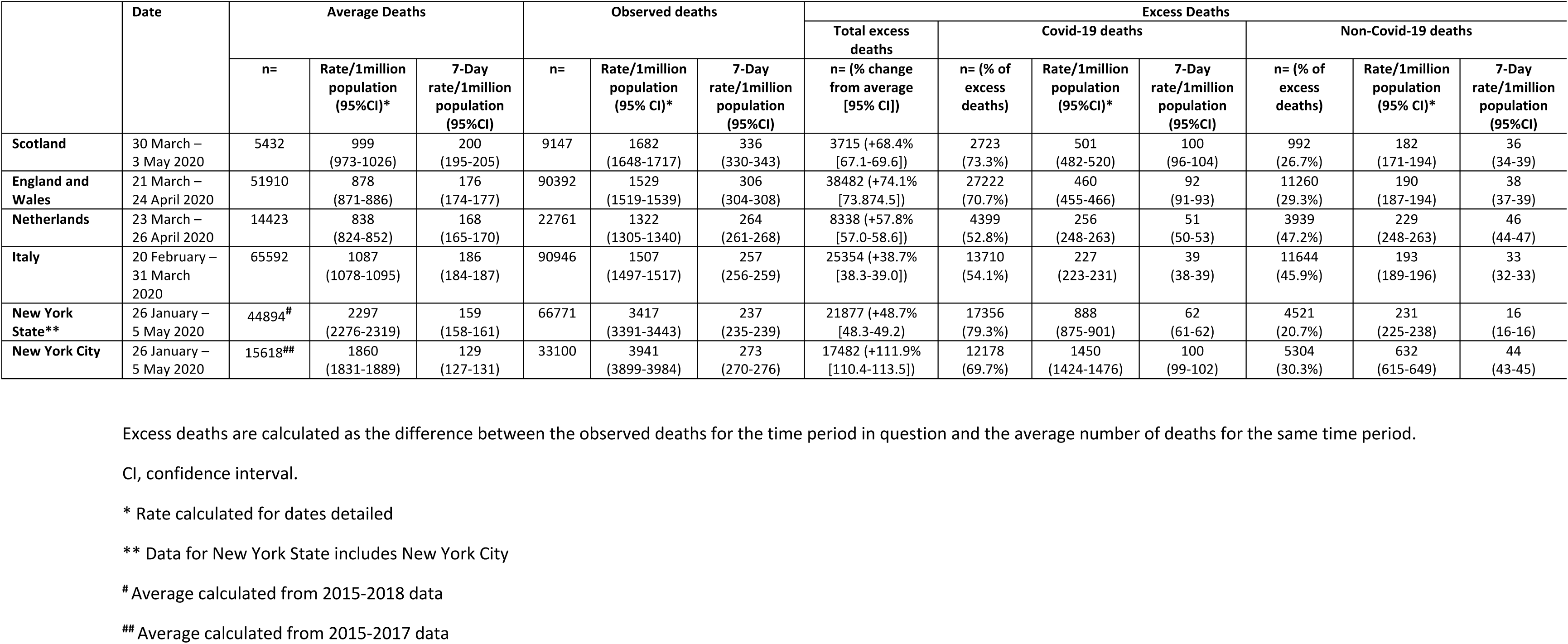
Deaths in Scotland, England and Wales, the Netherlands, Italy and New York State during the Covid-19 Pandemic.

